## Supplementary Figures for "Induction of trained immunity by influenza vaccination - impact on COVID-19"

**SUPPLEMENTARY MATERIALS**

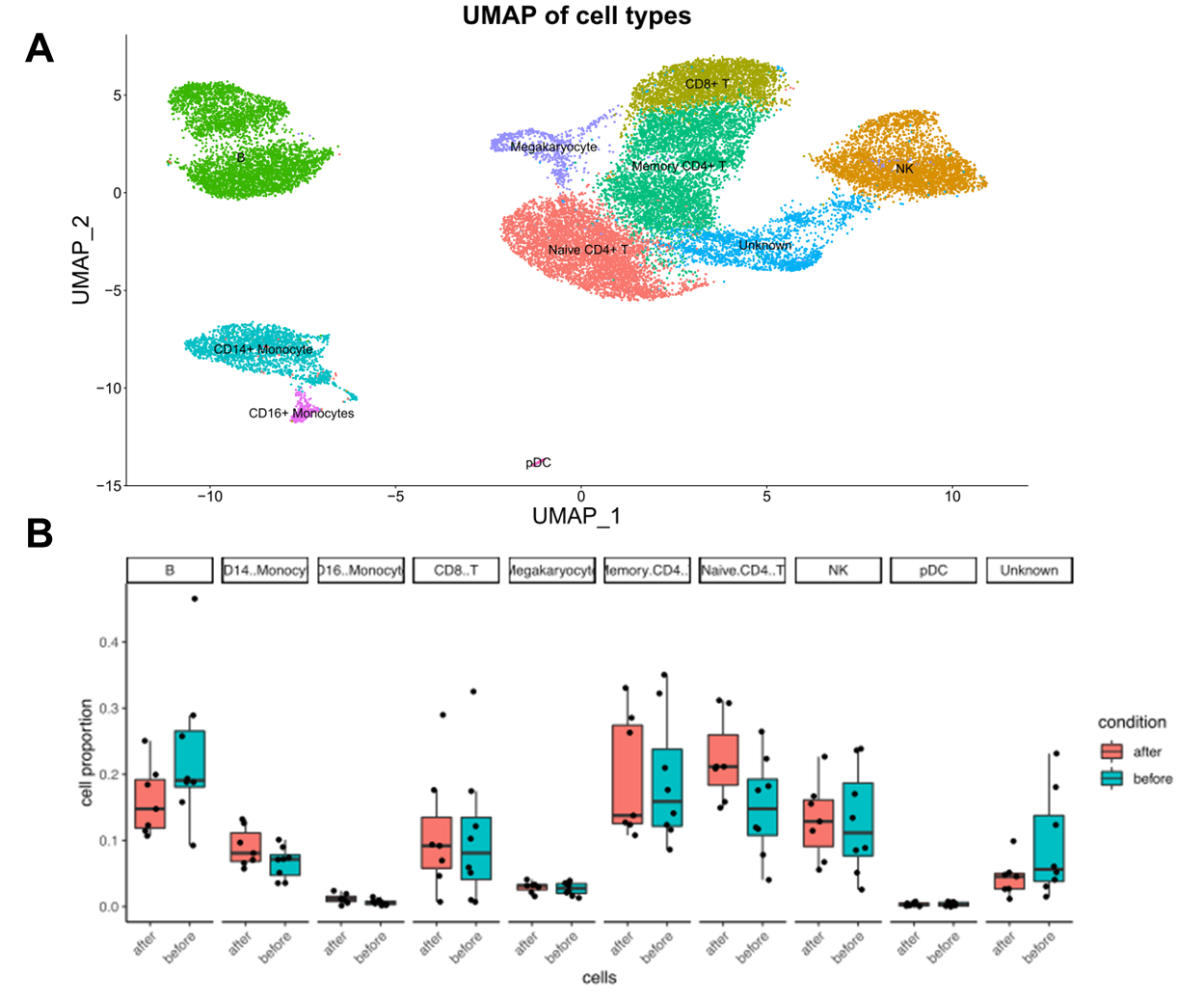

**S1 Figure: A.** Two-dimensional Uniform Manifold Approximation and Projection (UMAP) embedding of 25.562 single cells. Cells are colored respective to their major cell lineages. **B.** Proportions of immune cell populations 1 week before and 6 weeks after the influenza vaccination.

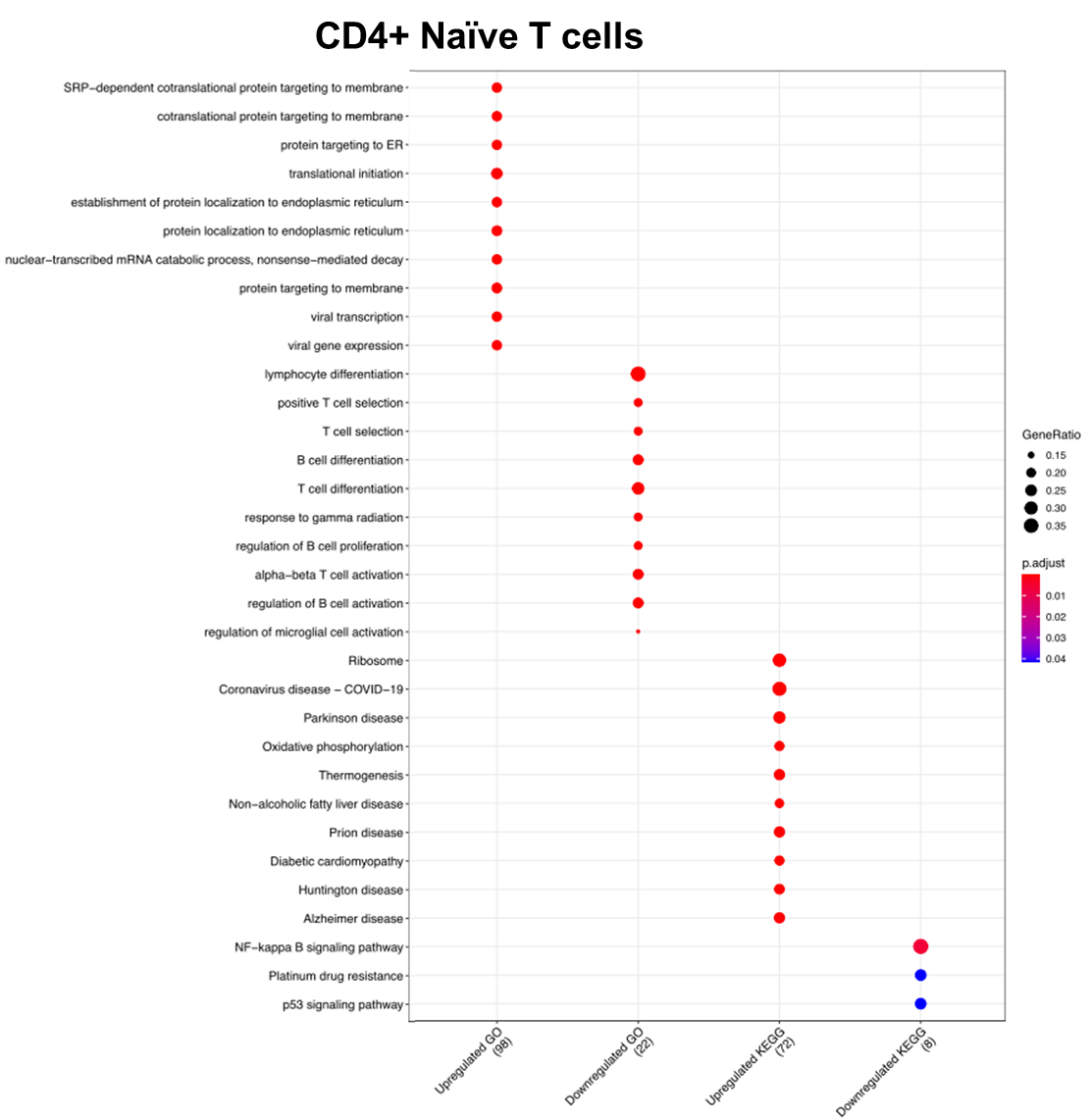

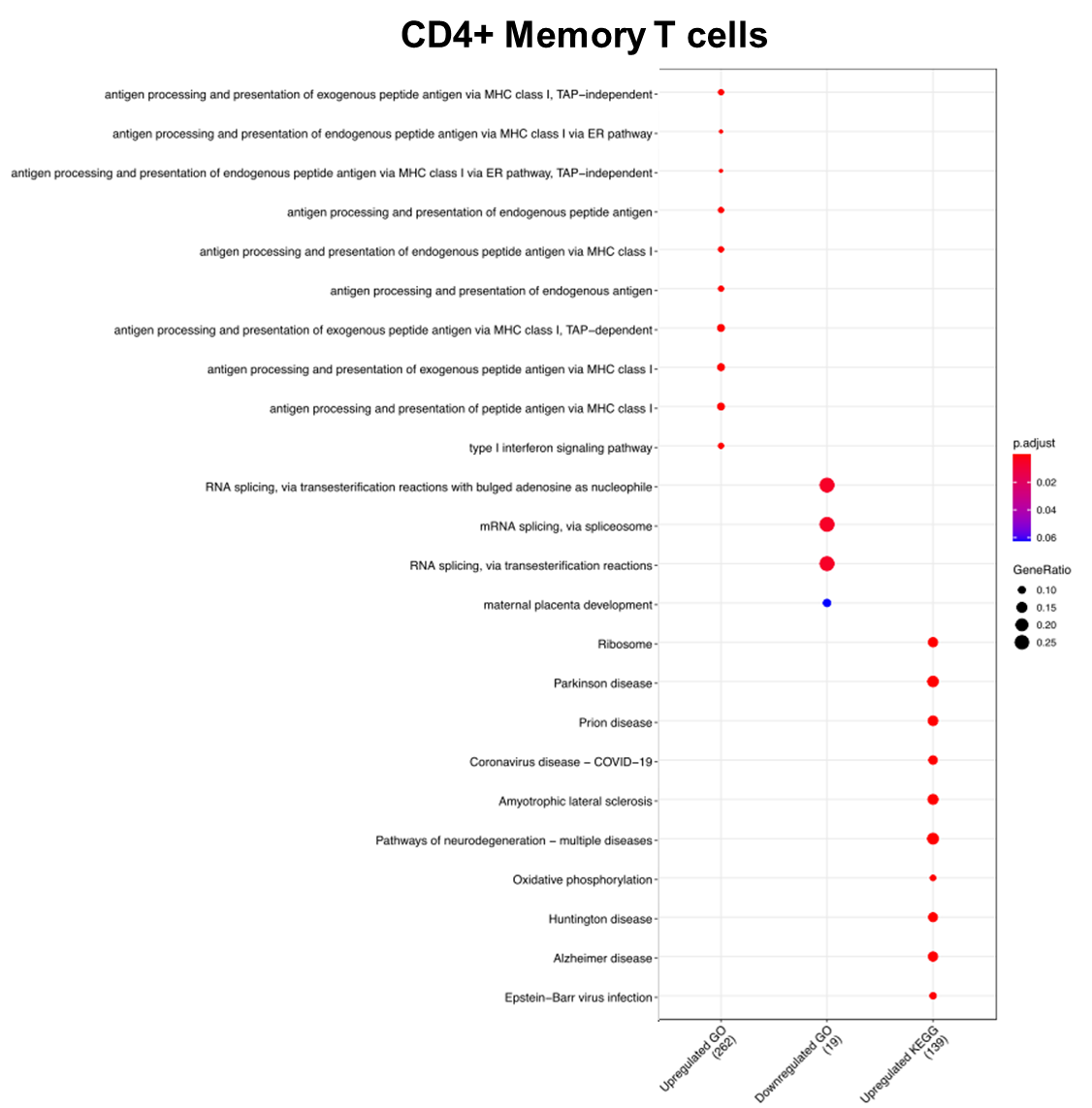

**S2 Figure: Gene ontology (GO) and Kyoto Encyclopedia of Genes and Genomes (KEGG) pathway enrichment analyses.** Analyses were performed with the genes whose expressions significantly change after vaccination in CD4+ naïve T cells (upper panel) and CD4+ memory T cells (lower panel).

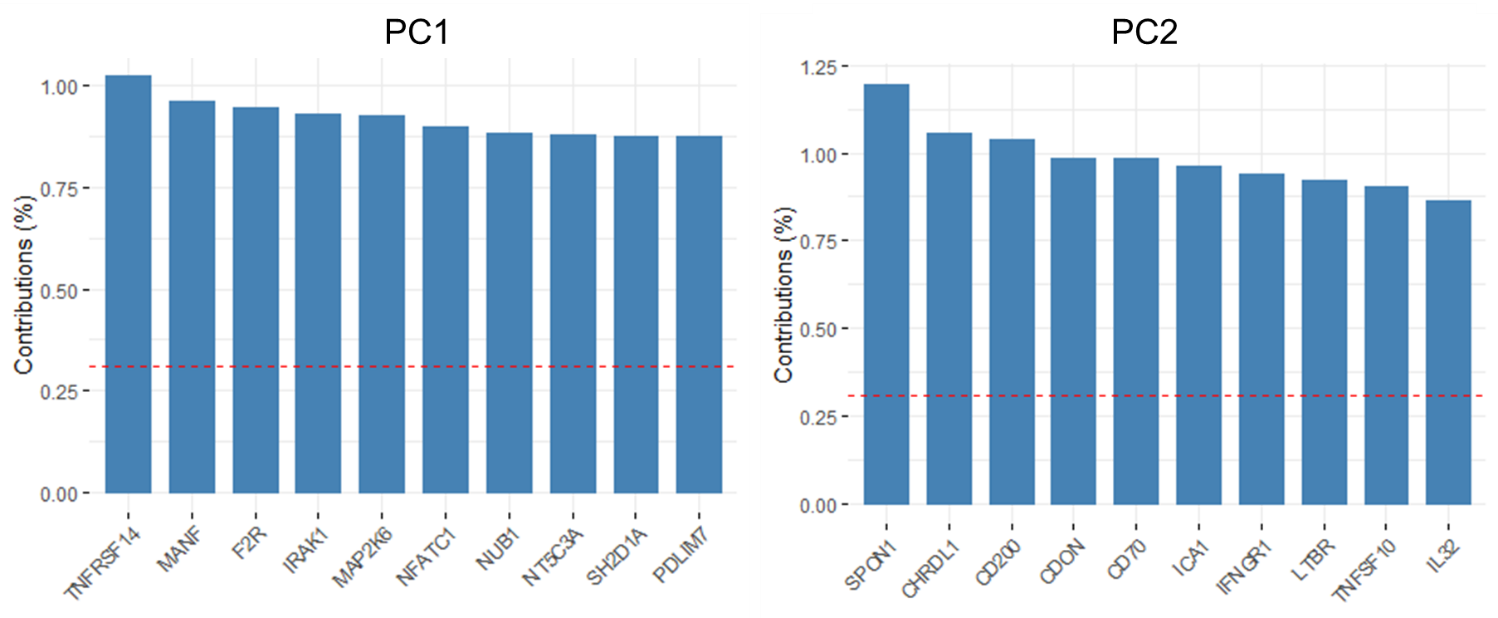

**S3 Figure: Contributions of individual proteins to the first two dimensions of the PCA performed with proteomics data before and after vaccination.** Left panel shows the proteins contributing to the variance in the first dimension (PC1), while right panel demonstrates the proteins contributing to the variance in the second dimension (PC2).

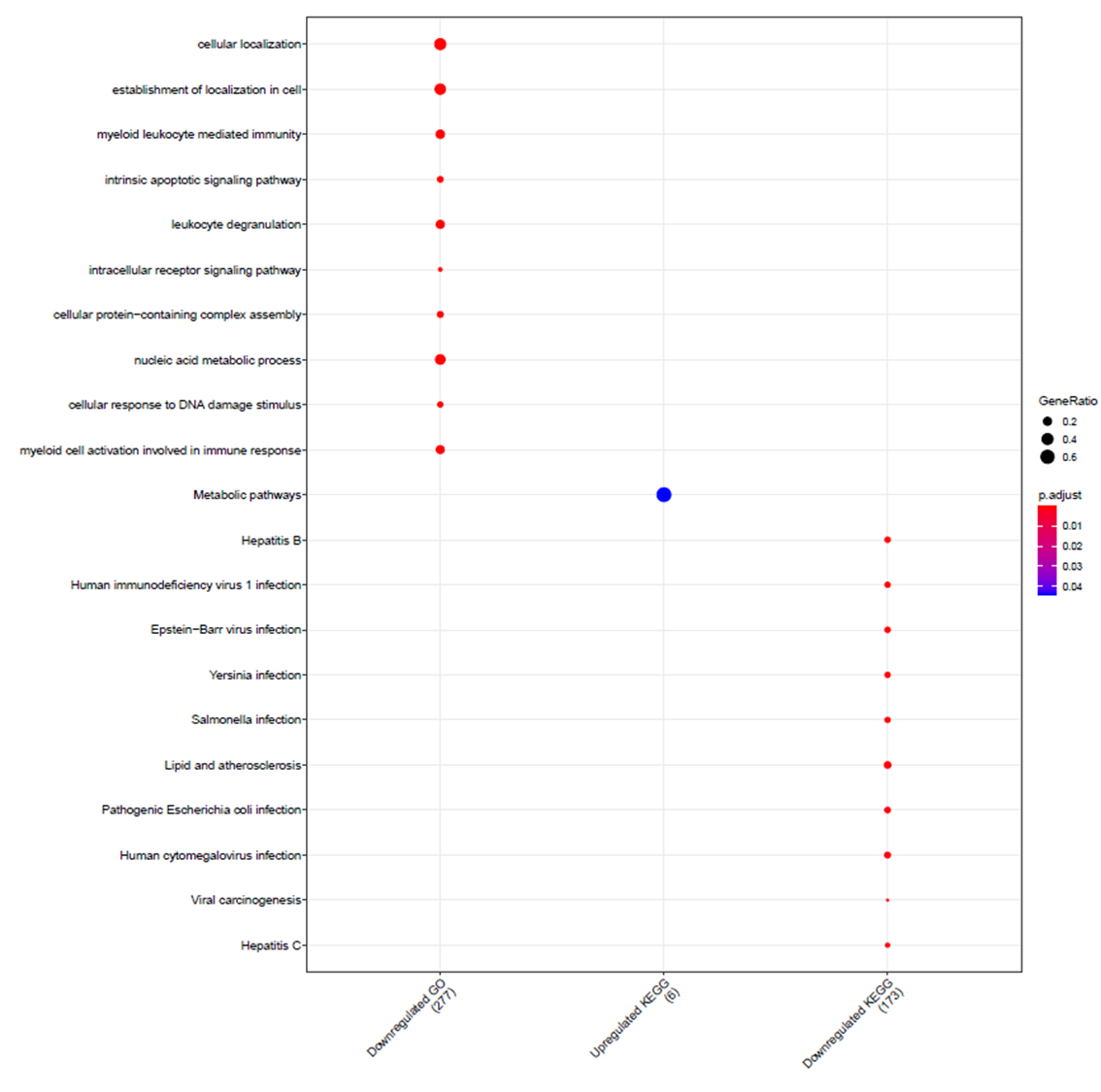

**S4 Figure. GO and KEGG pathway enrichment analyses using the proteins whose plasma concentrations significantly change after vaccination.** The complete Olink panel of 1472 proteins were used as the background in the enrichment analysis.

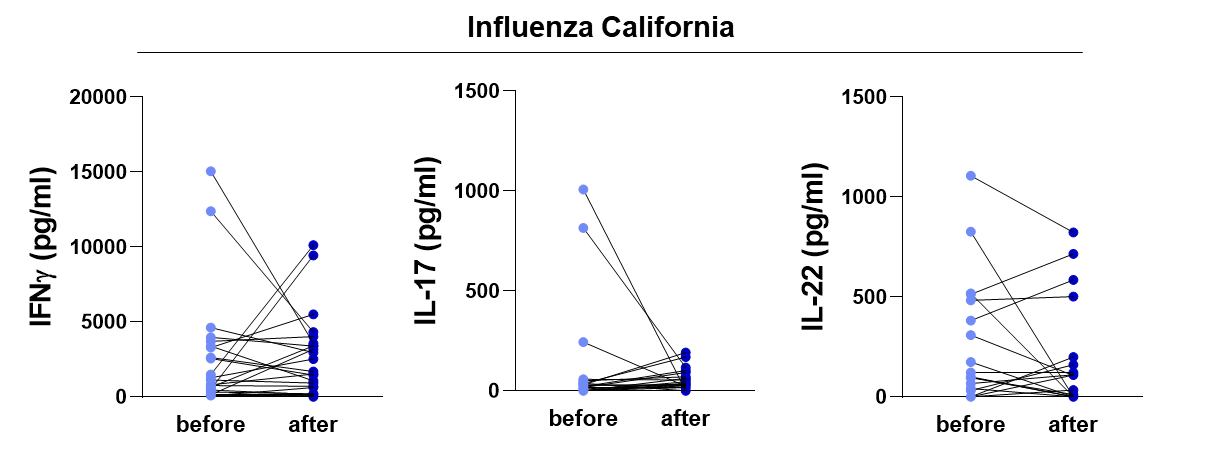

**S5 Figure:** **Ex vivo IFNγ, IL-17, and IL-22 responses of the individuals before and after influenza vaccination.** PBMCs were stimulated with heat-killed Influenza H1N1 (California strain) for 7 days. IFNγ, IL-17 and IL-22 responses were quantified. Wilcoxon signed-rank test revealed no significant differences in cytokine production between before and after influenza vaccination.
