## Supplementary figures and images for "Induction of trained immunity by influenza vaccination - impact on COVID-19"

### Supplementary Table

**S1 Table. Studies on the association between influenza vaccination and COVID-19 related outcomes.**

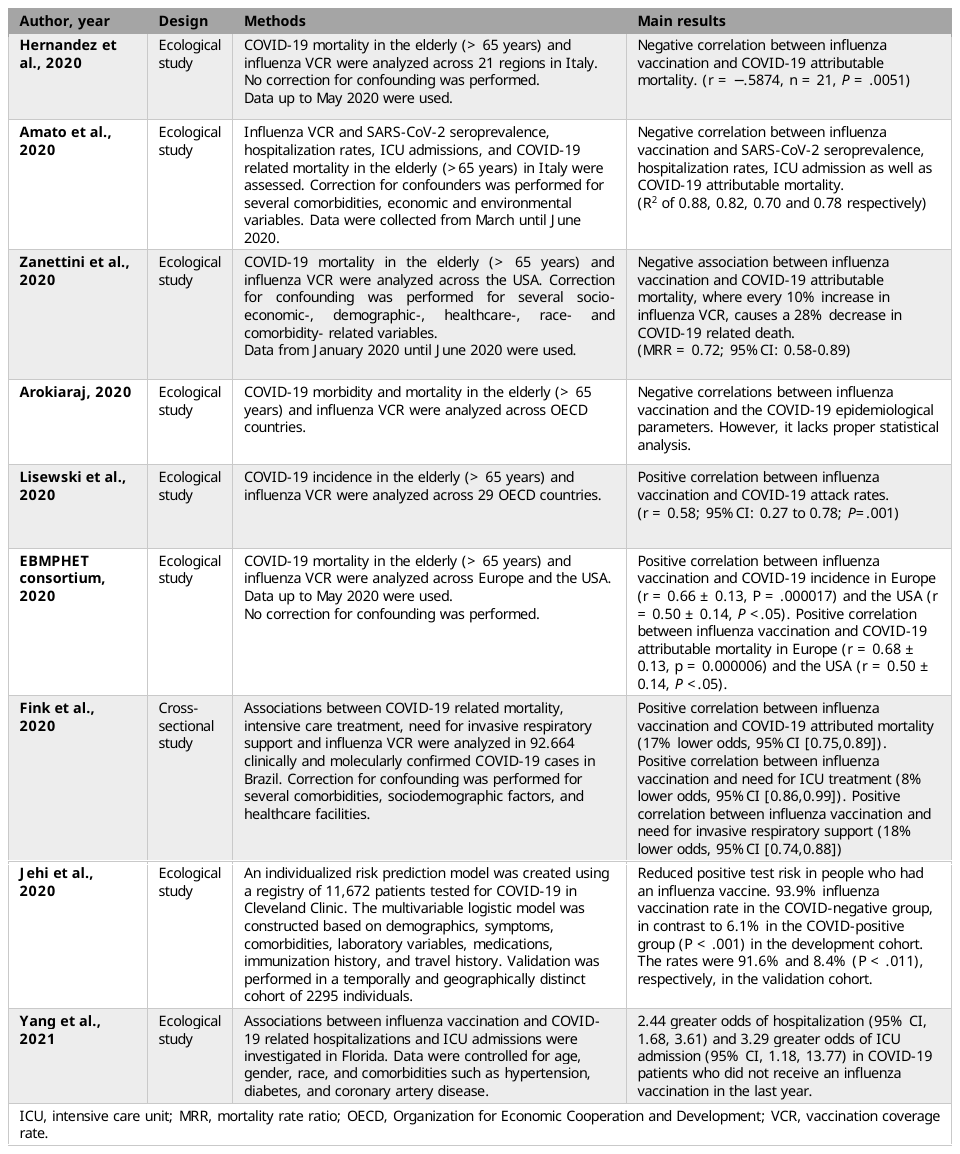
